# Detection of Cytomegalovirus infection in infants with biliary atresia: A meta-analysis

**DOI:** 10.1101/2020.05.04.20090134

**Authors:** Sagad Omer Obeid Mohamed, Almigdad H. M. Ali, Mohamed Elata Hassan Elbathani, Osman Omer Ali Abdallah, Assad Ahmed Mohammed Ahmed, Abazr A. H. Ibrahim, Almutasim B. E. Elhassan, Mohammed Suliman Tawer Salman, Mahmoud Elnil, Ibrahim H. E. Elkhidir, Mazin A.M. Elhassan, Abdulhamid Ibrahim Hassan Abuzied

**Affiliations:** Faculty of Medicine, University of Khartoum, Sudan

**Keywords:** *Cytomegalovirus*, Biliary atresia, prevalence, meta-analysis

## Abstract

**Background:** Biliary atresia (BA) is a leading cause of end-stage liver disease and the most common indication of liver transplantation in children. Several reports attributed BA to both prenatal and perinatal aetiologies, including a viral infection-induced auto-immune response that targets the bile ducts. *Cytomegalovirus* (CMV) remains the most common virus being linked to BA. This meta-analysis estimates to what extent CMV infection is detected in patients with BA.

**Methods:** This study was conducted according to the Preferred Reporting Items for Systematic Reviews and Meta-Analyses (PRISMA) guidelines. The databases of MEDLINE/PubMed, WHO-Virtual Health Library (VHL), ScienceDirect, and Google Scholar were used for the systematic search. Random effects model was used to estimate the pooled prevalence estimate with the corresponding 95% confidence interval (CI) using StatsDirect statistical software. Heterogeneity among the included studies was estimated using the I^2^ statistics. Publication bias was determined based on Begg’s test, Egger’s test, and examination of the funnel plot. Subgroup analyses and meta-regression were used to explore the moderators of heterogeneity between studies.

**Results:** A total of 13 studies that fulfilled the eligibility criteria were included in the meta-analysis. The pooled overall prevalence of CMV infection in patients with BA was 24.5% (95% CI 11.9 %– 33.9%). In subgroup and meta-regression analyses, the prevalence of CMV infection in patients with BA was correlated with the geographical areas (p = .001) but not with the study year (p = .153).

**Conclusions:** The available data demonstrate that the detection of CMV infection is common in infants with BA. There is still a need for large studies with appropriate controls to examine various aspects of the association between CMV infection and BA.

**Abbreviations:** *BA = biliary atresia, CMV = cytomegalovirus*.

## • Background

Biliary atresia (BA) is a frequent cause of neonatal cholestasis, characterized by extensive fibrosing inflammation of the extra-hepatic bile duct, leading to obstruction of bile flow, and subsequently resulting in permanent liver damage, if not early managed. [1–3]. BA is treated by Kasai portoenterostomy, which prevents liver injury caused by cholestasis if done early [1–2, 4]. Despite using Kasai operation, BA is still a leading cause of end-stage liver disease and the most common indication of liver transplantation in children [1, 4–6].

Biliary atresia is believed to be a disorder of multifactorial etiology, involving both prenatal and perinatal factors [1, 6]. Multiple studies support a possible infectious etiology, mostly, a viral infection-induced autoimmune response that targets the bile duct, leading to chronic fibro-sclerosing injury [1, 2, 4]. Patients with BA have been tested for several viruses in an attempt to determine the viruses associated with the disease onset. Among these viruses, *Cytomegalovirus* (CMV), remains the most common virus being linked to BA [5, 7].

Fischler et al. found that immunoglobulin deposits on the hepatocellular canalicular membrane were significantly higher in biopsies from CMV infected patients with BA than their controls [8]. Furthermore, histological findings in the porta hepatis samples of patients with BA supports the link between BA and a CMV infection [9].

Worldwide, Several studies assessed the prevalence of CMV infection in infants with BA. These studies gave different results of the prevalence of CMV infection in infants with biliary atresia, and to the best of our knowledge, there is no meta-analysis assessing the extent of CMV detection in biliary atresia patients. The systematic measurement and summary will show the magnitude of this problem and may aid in a better understanding of the disease etiology. This study aimed to estimate to what extent CMV infection is prevalent in patients with BA using a meta-analytic approach.

## • Methods

### Search strategy and inclusion criteria

In this study, we followed the guidelines of the Preferred Reporting Items for Systematic Reviews and Meta-Analyses (PRISMA) statement [10]. We searched the databases of Medline/PubMed, WHO-Virtual Health Library (VHL), Google Scholar, and ScienceDirect for all studies which assessed the association between CMV infection and BA up to September 2019. The search words used were “*Cytomegalovirus*,” “CMV,” and “Biliary atresia” to ensure the maximal coverage of possible literature.

Inclusion criteria: any study published in English that presented sufficient data for estimation of the prevalence of CMV infection in patients with BA was included. If two or more studies had the same patient population, the study with the more complete data or larger sample size was included to avoid duplication.

Case reports, case series, editorial letters, abstracts, and studies lacking the data of interest were excluded from this study.

Since BA is a congenital malformation, we included only studies that used a reliable method for diagnosing congenital CMV infection (viral DNA detection by PCR, viral isolation by culture, and viral antigen detection) [11–13]. Studies that relied only on serological testing were not included because the presence of CMV-IgG does not differentiate between infant infection and trans-placental transfer of maternal antibodies, and the presence of CMV-IgM is not sufficiently sensitive and does not differentiate between the congenital infection and acquired infection in the early neonatal period [11–12].

The titles and abstracts of all papers retrieved from this search were screened for potential inclusion in this review. Then, relevant studies were reviewed (full text) for inclusion according to the defined eligibility criteria, and any disparity between the reviewers was resolved by discussion and consensus. Quality of the studies was assessed using the Newcastle – Ottawa scale, a tool that determines the quality based on the selection of the study group, comparability of groups, and ascertainment of the exposure and outcomes. Data were extracted using a data extraction form developed to extract the following data: authors, study region, year of publication, diagnostic methods used for detection of CMV infection, number of patients with BA, and number of patients with confirmed CMV infection.

### Statistical analysis

The pooled prevalence from the random-effects models with the corresponding 95% confidence interval (CI) was calculated using StatsDirect statistical software version 3.2.8 for analysis. Heterogeneity among studies was estimated using the I^2^ statistics, and publication bias was estimated through visual examination of the funnel plot, Begg’s test, and Egger’s test [14–15]. Chisquare (X^2^) test was used to assess the differences between the categorical subgroups, and the significance level was set at 0.05. We conducted subgroup analyses and meta-regression to determine the extent to which variables of interest moderated the overall result.

## • Results

The schematic flow of study identification and selection process is presented in (Figure 1). The initial search retrieved records for 401 published articles. We excluded 365 articles that were obviously irrelevant or duplicated in the databases. The remaining 36 studies were retrieved for a full-text assessment, and 23 studies were subsequently omitted because of low quality and lack of sufficient data for estimation of the outcomes of interest. Lastly, a total of 13 studies published from 1980 to 2018 which met the eligibility for data extraction and analyses were used for the quantitative synthesis; six studies from Asia [16–21], three studies from Europe [22–24], three studies from the Americas [25–27], and a study from Africa [28]. The main characteristics of these included studies are shown in (Table 1).

**Figure 1:**
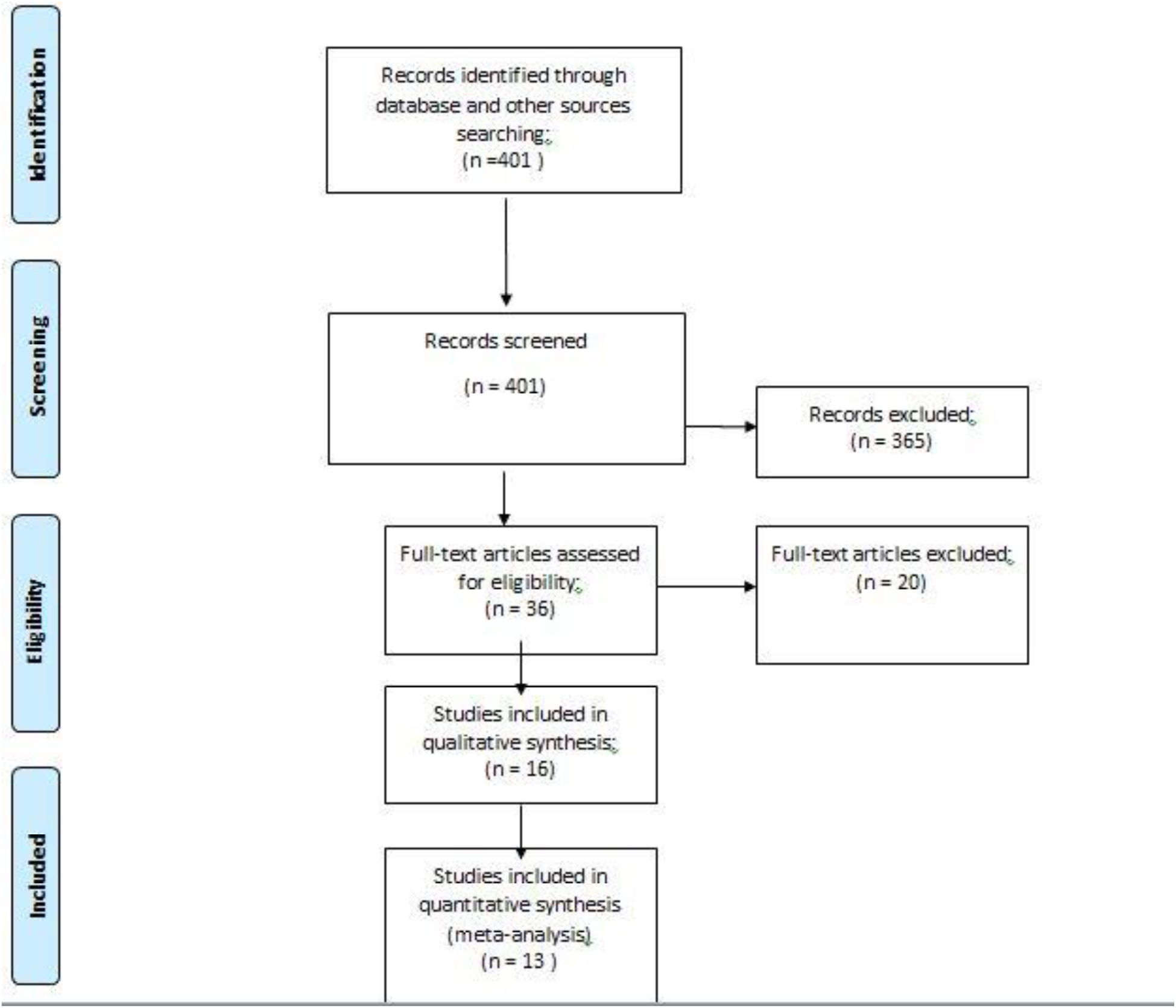
The flow diagram for the process of study selection and systematic review of literature.

**Table 1:** Descriptive summary of the studies included in the review.

| First Author | Year | Country | Detection of CMV | No. of patients with BA | No. of cases with CMV |
| --- | --- | --- | --- | --- | --- |
| Xu et al. | 2011 | China | PCR | 85 | 51 |
| Shen, et al. | 2008 | China | Antigen detection | 27 | 10 |
| Soomro, et al. | 2011 | Pakistan | PCR | 33 | 14 |
| Yaghobi, et al. | 2010 | Iran | PCR | 34 | 0 |
| Goel, et al. | 2018 | India | PCR | 14 | 11 |
| Numazaki et al | 1980 | Japan | Viral isolation (culture) | 18 | 2 |
| Fischler, et al | 1998 | Sweden | PCR | 18 | 9 |
| Schukfeh, et al. | 2012 | Germany | PCR | 70 | 8 |
| Fjær, et al. | 2005 | Norway | PCR | 10 | 2 |
| Tarr et al. | 1996 | USA | Viral isolation (culture) | 23 | 4 |
| Jevon, et al. | 1999 | Canada | PCR | 12 | 0 |
| De-Tommaso, et al | 2015 | Brazil | PCR | 33 | 9 |
| Goedhals, et al. | 2008 | South Africa | Antigen detection | 42 | 1 |

Meta-analysis showed that CMV was detected in 24.5% (95% CI 11.9%–33.9%) of the patients with BA (Figure 2). Slight publication bias was detected on visual examination of the funnel plot (Figure 3) and from the results of Begg’s test (p = 0.03) and Egger’s test (p = 0.01). In subgroup analysis, the detection of CMV infection in patients with BA was higher in the Asian studies (37.9%) than in European and American studies (25.5% and 15.3%, respectively). There was a statistical difference in prevalence between the aforementioned geographical areas (X^2^ = 21.54, P <0.001). A meta-regression analysis was done to analyze whether the study period affected the heterogeneity among studies in this meta-analysis, and the results showed that the years were not correlated with the outcome (coefficient 0.009: P = 0.152).

**Figure 2:**
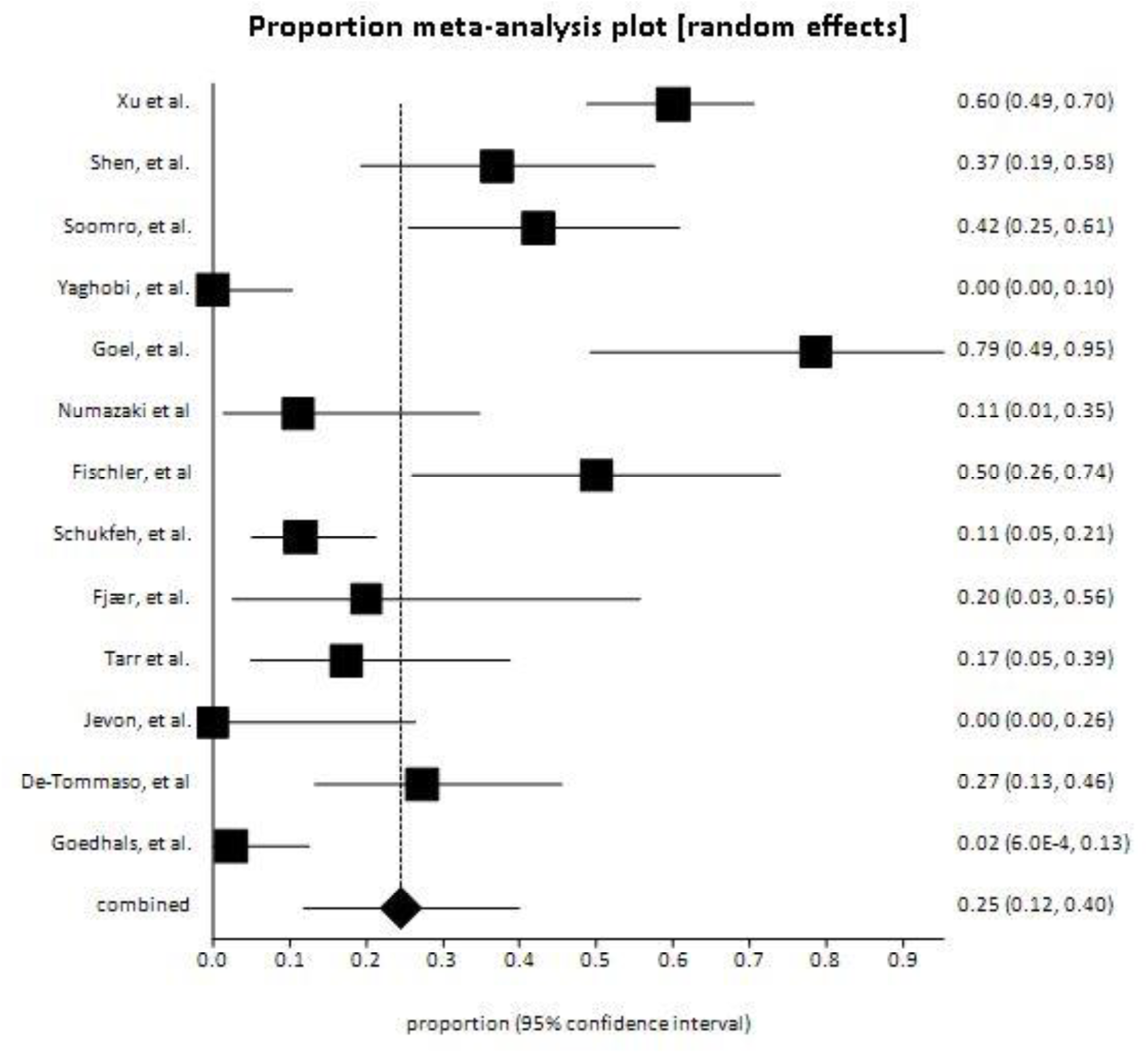
Pooled prevalence of CMV infection among patients with BA.

**Figure 3:**
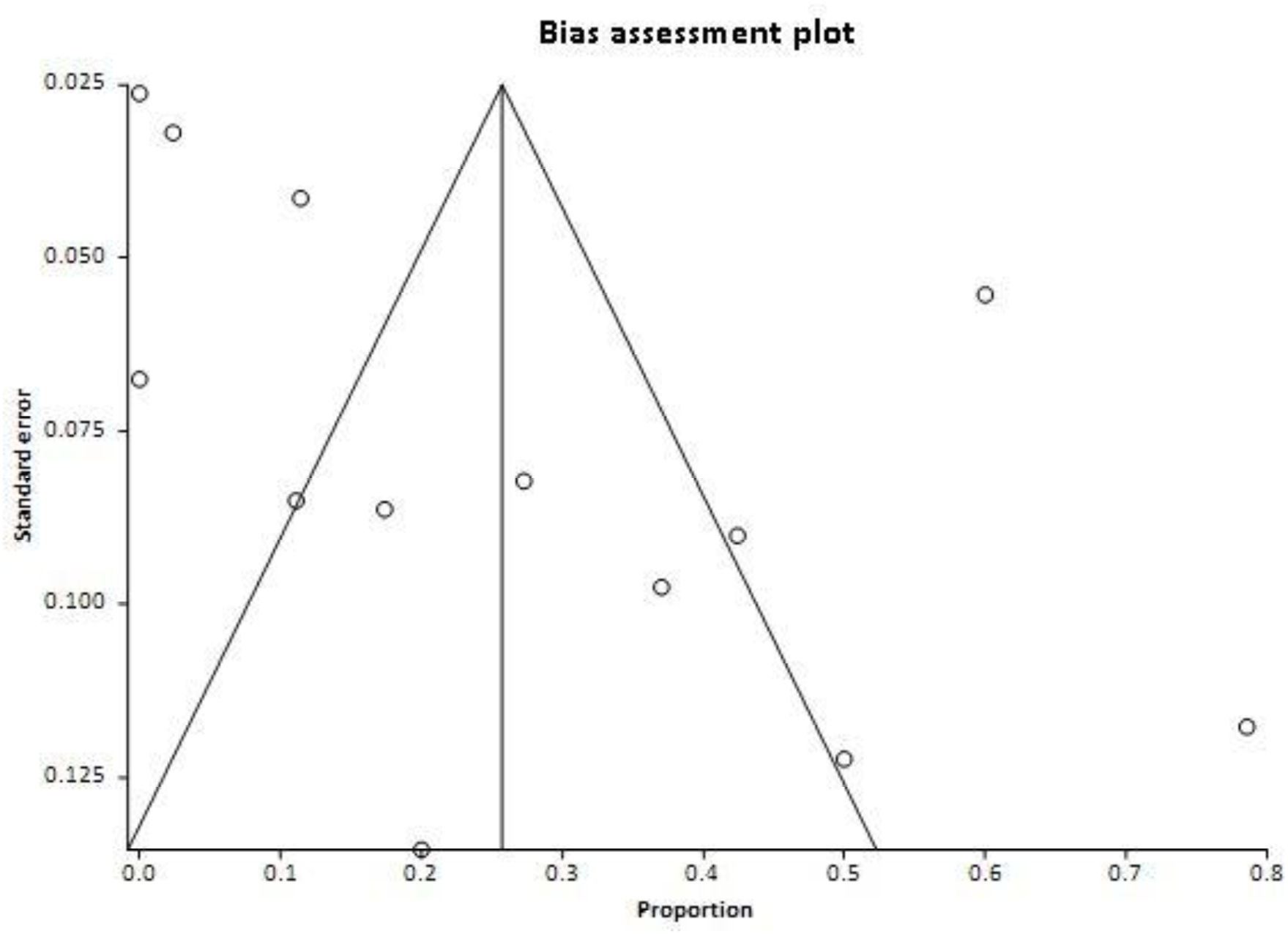
Funnel plot for assessment of the publication bias.

## • Discussion

CMV is a well-known cause of congenital viral infections in humans. It has been proposed as a possible etiologic agent in the pathogenesis of BA. In a meta-analysis conducted in 2007, it was reported that the overall birth prevalence of congenital CMV infection in the general population was only 0.64%, and the majority of the cases were asymptomatic [29]. However, this meta-analysis showed a remarkably higher prevalence of CMV infection in infants with BA (24.5%) than that of congenital CMV infection in the general population.

The studies included in this meta-analysis were mostly from the Americas, Asia, and Europe, and only one study from Africa. Though there is limited literature published on the association between CMV infection and BA, the available data demonstrate that CMV infection is common in BA as around one-quarter of the infants with BA had evidence of CMV infection using one of the reliable methods of CMV detection (viral DNA detection by PCR, viral isolation by culture, and viral antigen detection).

The discrepancy between prevalence rates reported from the reviewed studies might be partially explained by the differences in sensitivity of the detection method used or by the presence of other etiological agents implicated in the pathogenesis of BA. In this meta-analysis, CMV infection in patients with BA showed regional epidemiological differences. While rates of CMV infection have been reported to be considerably lower in most European and American countries, the prevalence rate tends to be higher in Asian countries. This finding is consistent with a socioeconomic link with CMV that has been well established in several studies, showing that CMV prevalence is higher in individuals of areas of lower socioeconomic groups [30]. Furthermore, some studies demonstrate that BA is more prevalent in Asia than in Europe. It occurs in 1/5 - 1/8,000 live births in Asia and 1/18 - 1/20,000 children in Europe, although no ethnic differences have been demonstrated [26, 35]. Also, the regional differences in times of CMV epidemics could aid in the understanding of this variation [5].

Davenport et al. have classified BA into four groups: isolated BA, cystic BA, syndrome BA with associated malformation, and CMV-associated BA [31]. Among these types of BA, it has been reported that the CMV-associated BA yielded the worst prognosis [17, 32]. Shen et al. reported that the CMV infection group had a lower rate of jaundice disappearance and a higher incidence of postoperative ascending cholangitis, a significant complication of BA [17]. Zani et al. findings suggest that CMV-associated BA is a distinct etiological and prognostic subgroup, with reduced clearance of jaundice, native liver survival, diminished response to Kasai portoenterostomy, and increased mortality [33]. Unfortunately, Because of the lack of uniformity across the included studies, a systematic assessment of the prognosis of CMV-associated BA couldn’t be done.

Another important concern claimed by several authors regarding the association between CMV infection and BA is that detection of CMV infection might confound the clinical evaluation of neonatal jaundice caused by BA and may lead to misdiagnosis, late referral, and a delay in therapeutic surgery. This usually occurs when the condition is misdiagnosed as neonatal hepatitis secondary to CMV infection [28, 27, 32].

There is a suggested role of antivirals in the treatment of BA patients infected with CMV. It could be a possible intervention and may consequently improve outcomes of BA [17, 27]. A recent trial done by Parolini et al. showed that antiviral treatment with valganciclovir and ganciclovir appeared to improve outcomes in infants with CMV-associated BA [34]. However, there is a need for more trials to confirm the effect of antiviral treatment among those patients.

## • Conclusions

This study showed that CMV infection was detected in 24.5% of the infants with BA. There is still a need for large prospective multicenter studies with appropriate controls to examine various aspects of the association between CMV infection and BA.

## Data Availability

The dataset generated during this study are available from the corresponding author on reasonable request.

## Declarations

- **Ethical approval and consent to participate:** not applicable
- **Consent for publication:** not applicable.
- **Availability of data and material:** The dataset generated during this study are available from the corresponding author on reasonable request.
- **Competing interests:** The authors declare that they have no competing interests.
- **Funding:** No fund
- **Authors contribution:** (SM) conceptualized the research idea and designed the study; (SM, AA, ME and AI) undertook articles searching, articles assessment, and review; (SM, ME, and AE) undertook data extraction and analysis; All authors interpreted the results and drafted the manuscript. All authors revised and approved the final manuscript.
- **Acknowledgment** Non to acknowledge

## Notes

### Competing Interest Statement

The authors have declared no competing interest.

